# REcovery and SURvival of patients with moderate to severe acute REspiratory distress syndrome (ARDS) due to COVID-19: a multicentre, single-arm, Phase IV Itolizumab Trial: RESURRECT

**DOI:** 10.1101/2021.10.25.21265462

**Authors:** KR Raveendra, Chirag Rathod, Rahul Darnule, Subramanian Loganathan, Sarika Deodhar, A Radhika, Ashwani Marwah, Nitin M Chaudhari, Binay K Thakur, Sivakumar Vaidyanathan, Sandeep Nilkanth Athalye

## Abstract

**Objective:** To evaluate safety and efficacy of Itolizumab in hospitalized COVID-19 patients with PaO_2_/FiO_2_ ratio (PFR) ≤200 requiring oxygen therapy.

**Design:** A multicentre, single-arm, Phase-4 study with a treatment period of 30-Days and an extended follow-up period of 90-Days.

**Methods:** Hospitalized adult patients (n=300) with SARS-CoV-2 infection, with PFR ≤200; oxygen saturation ≤94% and ≥1 elevated inflammatory markers were included from 17 COVID-19-specific tertiary hospitals in India. Patients received Itolizumab infusion 1.6 mg/kg and were assessed for 1-month and then followed up to Day-90.

**Results:** Day-30 post-treatment safety/efficacy results and Day-90 mortality results are presented. Primary outcome measures: incidence of severe acute infusion-related reactions (IRRs) (≥Grade-3) was 1.3% and mortality rate at 1-month was 6.7% (n=20/300). Key secondary analyses: Mortality rate at Day-90 was 8.0% (24/300). 91.7% patients came off the oxygen therapy within Day-30 of treatment. By Day-7, most patients had stable/improved SpO_2_ without increasing FiO_2_. Mean PFR improved by 50% by Day-7 (p<0.001) and the trend remained consistent till Day-30. Median time of recovery was 8 days. Cumulatively, at Day-30, 260(86.7%), 256(85.3%), 132(44.0%), 113(37.6%) and 32(10.7%) patients showed >1-point, >2-point, >3-point, >4-point and 5-point improvement on the modified COVID-19 8-point ordinal scale and worsening of symptoms by >1 point, >2 points and 3-points was seen in 26(8.7%), 20(6.7%) and 6(2.0%) patients, respectively. CRP, D-dimer, LDH, and serum ferritin levels significantly decreased (p≤0.01) compared with baseline. IL-6 and TNFα levels also decreased 48-hours post-infusion. Overall, 123 treatment-emergent adverse events (TEAEs) were reported in 63 patients, most being Grades 1-3. Most common TEAEs were IRRs and lymphopenia; most common serious TEAEs were septic shock, worsening of ARDS, and respiratory failure. No deaths were attributable to Itolizumab.

**Conclusion:** Itolizumab shows no new safety concerns and suggests a mortality and recovery benefit at 1-month in hospitalized COVID-19 patients requiring oxygen therapy.

**Trial registry number:** CTRI/2020/09/027941

## Introduction

The coronavirus pandemic caused by SARS-CoV-2 (COVID-19) infection has, as of date, spread across the globe, with ∼250 million confirmed cases and ∼5 million deaths.^1^ Patients with an increasing severity of the disease can develop hypoxaemia/dyspnoea, and can potentially evolve towards an acute respiratory distress syndrome (ARDS) due to an exaggerated immune response. This stage of the disease is characterized by high levels of pro-inflammatory cytokines such as interferon-γ, interleukin-6 (IL-6), IL-1β, IL-17, IL-18, tumour necrosis factor-alpha (TNF-α), granulocyte-macrophage colony-stimulating factor etc., and without a timely and urgent intervention may progress towards death.^2-4^ There is an elevation of other markers of inflammation, coagulation and organ damage such as C-reactive protein (CRP), D-dimer, lactate dehydrogenase (LDH) and ferritin.^5^ Treatment with immunomodulatory drugs aiming to reduce elevated cytokines have shown survival benefits in patients with hyperinflammation, without an increase in adverse events (AEs).^6-9^

Itolizumab is a humanized IgG1 monoclonal antibody that binds to domain-1 of human-CD6 responsible for priming, activation and differentiation of T-cells.^10^ By selectively targeting the CD6-ALCAM pathway, Itolizumab results in decreased IFN-γ, IL-6 and TNF-α levels through the Th-1 pathway and IL-17, IL-6 and TNFα levels through the Th-17 pathway.^10-16^ Itolizumab leads to a reduction in T-cell infiltration at the sites of inflammation without any T-cell depletion.^10^ Itolizumab was granted a Restricted Emergency Use Authorization (EUA) in India for the treatment of moderate-to-severe ARDS in patients with COVID-19 infection, based on the favourable outcomes from a smaller controlled trial.^9^ In the past 1 year, >27,000 hospitalized patients have been treated for COVID-19 with Itolizumab across India.^17-23^

This post-marketing study in 300 patients, hospitalized in COVID-19-designated Indian tertiary care hospitals, was carried out to assess safety and survival and recovery benefits of Itolizumab treatment in COVID-19 patients requiring oxygen therapy and having a PaO_2_/FiO_2_ ratio (PFR) ≤200. This study presents results of the primary and key secondary analyses of efficacy and safety endpoints for a 30-day post-treatment period with Itolizumab. The study was designed for an extended follow-up period up to 90 days after treatment.

## Methods

### Study design

This was a multicentre, single-arm, Phase-4 trial where 300 eligible patients in India were dosed with Itolizumab, as per the prescribing information, in addition to receiving the best supportive care (BSC; ***Suppl. Figure 1***). Patients were observed for 30 days for mortality and were followed up to 90 days post-treatment for long-term survival as well as any benefit of Itolizumab treatment in long-term complications of COVID-19. The study flow chart is summarized in **Figure 1**. This publication presents the 30-day safety and efficacy data and the 90-day mortality rate.

**Figure 1:**
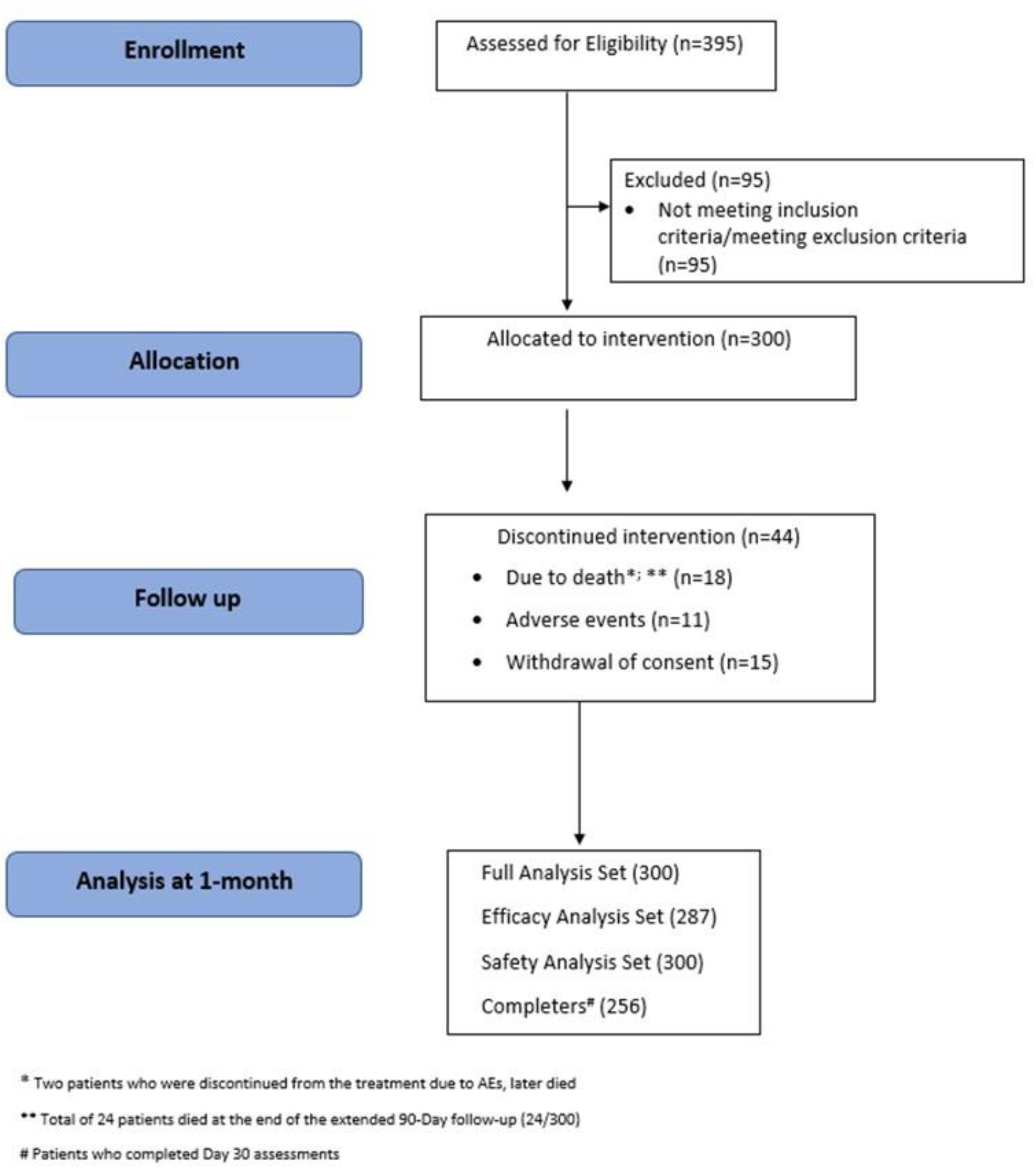
The study flow chart.

### Study participants

The following patients were included in the study: Adult male or female patients with confirmed virologic diagnosis of SARS-CoV-2 infection (reverse transcriptase-polymerase chain reaction/rapid antigen test/equivalent), hospitalized with moderate-to-severe ARDS due to worsening of COVID-19 as defined by PFR ratio of ≤200 with oxygen saturation at ≤94% at rest in ambient air or requiring supplemental oxygen therapy with one or more inflammatory markers (serum ferritin, LDH, D-dimer, CRP or IL-6) raised above the upper limit of normal.

Patients with known severe allergic reactions to monoclonal antibodies, with active tuberculosis (TB)/inadequately treated TB/latent TB, on oral anti-rejection or any immune-suppressive drug in the last 6 months and those who had participated in any clinical trial using an anti-IL-6 therapy, were excluded from the study. Patients with a known history of Hepatitis B, C or HIV, absolute neutrophil count <1000/mm^3^, platelet count <50,000/mm^3^ and absolute lymphocyte count <500/mm^3^ were also excluded. Patients on invasive mechanical ventilation (IMV), in whom progression to IMV or with high probability of death in the next 24-hours were also excluded. The study was carried out at 17 COVID-19-specific tertiary hospitals in India between 02 Oct 2020 and 10 Jul 2021.

### Procedures and Assessments

A modified COVID-19, 8-point ordinal scale was used to characterize the severity of the disease in the patients at baseline ***(Suppl. Table 1)***.^24^ Itolizumab infusion was initiated after a pre-medication with Hydrocortisone 100 mg i.v. and Pheniramine 30 mg i.v. about 30±10 minutes before the infusion. Patients were initiated on 1.6 mg/kg i.v. infusion of Itolizumab and one additional dose of 0.8 mg/kg was administered 8 days later, in patients with clinical deterioration and increase in serum inflammatory markers, at the investigator’s discretion.

Patients with treatment-emergent AEs (TEAEs) were assessed during the treatment and up to Day-30 as per Common Terminology Criteria for Adverse Event (CTCAE) version 5.0. Vital signs, other physical examinations and laboratory tests were carried out as per schedule. Blood samples were collected at scheduled time points for analysis of cytokines (IL-6 and TNF-α) and inflammatory markers (CRP levels, serum ferritin, D-dimer and LDH). A full description of assessments is available in the ***Supplementary Appendix***.

This study was carried out following the ethical principles described in the Declaration of Helsinki (64th WMA General Assembly, Fortaleza, Brazil, October 2013), the International Council for Harmonization Good Clinical Practice (ICH GCP) E6 (R2), New Drugs and Clinical Trials Rule-2019 issued by the Government of India and ethical guidelines for biomedical research on human subjects issued by the Indian Council of Medical Research. The study received approvals from the Independent Ethics Committees of all the participating sites. The patients provided written informed consent before the initiation of study procedures. The trial protocol was registered with the Clinical Trials Registry of India (CTRI/2020/09/027941).

### Study Objectives and Endpoints

The current study was undertaken with the primary objective of evaluating the safety and efficacy of Itolizumab infusion in patients with moderate-to-severe ARDS due to COVID-19. The secondary objectives of the study were to evaluate the additional efficacy and safety of Itolizumab as per the study outcomes detailed for the first 30 days out of the 90 days extended follow-up after the Itolizumab treatment.

The study included two primary outcome measures (1) safety-related outcome measure, namely, the incidence of severe, acute infusion-related reactions (IRRs), and higher, occurring on the day of infusion and (2) mortality rate at 1-month. The key secondary outcome measures included measurement of biomarkers (cytokines IL-6 and TNF-α) and inflammatory markers (CRP, serum ferritin, D-dimer and LDH). Other key secondary measures included measurement of lung function (stable/improved SpO_2_ without increasing FiO_2_ and PaO_2_ [partial pressure of oxygen]/FiO_2_ [fraction of inspired oxygen] ratio [PFR]), duration of hospitalization and intensive care unit (ICU) stay, remission of respiratory symptoms, improvement of the clinical status by 1-2 points on the modified COVID-19 8-point ordinal scale and safety.

### Statistical Analysis

Continuous variables were summarized using descriptive statistics such as mean, standard deviation (SD), median, 95% confidence interval (CI), minimum and maximum. Categorical variables were summarized using proportions (counts and percentages). Data were summarized at screening/baseline, 24-48 hours post-dosing, at Day-7, at the day of discharge and end of study at Day-30.

The primary safety endpoint was incidence of severe acute IRRs and higher as assessed by CTCAE. This primary endpoint was descriptive in nature as no hypothesis was specified. Summary statistics (frequency, percentage, point estimable, 95% CI using Clopper–Pearson exact test) of incidence of nature and severity of AEs that were severe acute IRRs, and higher, was calculated for the safety analysis set.

The 1-month mortality rate, which was the primary efficacy endpoint, was descriptive in nature as no hypothesis was specified. Summary statistics (frequency, percentage, point estimable, 95% CI using Clopper–Pearson exact test) was calculated for the full analysis set and efficacy analysis set. The mortality rate at Day-30 and time to death were summarized with descriptive statistics.

All safety analyses were conducted using the safety analysis set. AEs were coded using the Medical Dictionary for Regulatory Activities (MedDRA, version 23.1). The number and percentage of patients with AEs were tabulated by system organ class/preferred term and displayed by intensity and relationship to the study drug. Vital signs and laboratory data were summarized by patient.

The Full Analysis Set (FAS) included all patients who were enrolled in the study (ITT population). Efficacy Analysis Set included all patients of the FAS population who received at-least one full infusion of Itolizumab (without any major protocol deviation; decided before the database lock) and was used for efficacy endpoint analysis (modified ITT [mITT] population). The Safety Analysis Set included all patients who received a partial or complete infusion of Itolizumab, and this was used for all safety endpoint analysis (safety population).

All statistical tests were performed at the 5% level of significance (two-sided test) and p ≤0.05 was considered statistically significant. All statistical analyses were performed using SAS^®^ (version 9.4 or higher) system software (SAS Institute Inc., USA/R/EAST/NCSS software).

## Results

### Participant disposition and baseline characteristics

A total of 395 patients were screened, of which 95 were considered screen failures. The ITT population comprised of 300 patients and the mITT population comprised of 287 (95.7%) patients. A total of 44 patients discontinued from the study and the primary reason was death (18, 6.0%), adverse events (11, 3.7%), or withdrawal of consent (15, 5.0%). Additionally, two patients who were discontinued from the treatment due to AEs died later, resulting in a 1-month mortality rate of 6.7% (20/300). At the end of the 90-Day follow-up period, a total of 24 deaths (8%; 24/300) were reported.

Patients requiring oxygen therapy at enrolment were on non-invasive ventilation (NIV: bi-level positive airway pressure), high flow nasal cannula (HFNC), non-rebreather mask, a face mask or nasal prongs ***(Suppl. Table 1)***. Of the total 300 enrolled patients, 74.0% (n=222) patients were at score 5 (requiring any supplemental oxygen) and 26.0% (n=78) were on score 6 (requiring NIV or use of HFNC) on the modified ordinal scale ***(Suppl. Table 2)***.

Thirteen patients (4.3%) could not complete the infusion mainly due to IRRs. Overall, 9 patients (3.0%) required a second discretionary dose of Itolizumab infusion upon assessment by the investigator. ***Suppl. Table 3*** mentions the number of Itolizumab doses provided to each patient. The average duration of each Itolizumab infusion was 5-6 hours. All IRRs resolved on the same day, including in patients where the infusions were interrupted.

The mean age of patients in the trial was 53.3 years and all patients were of Asian ethnicity, with 74.7% of the patients being male. The disposition, demographics and baseline characteristics of the patients are given in ***Suppl. Table 4***. One hundred and fifty patients (150; 50%) had at least one medical condition, with hypertension as the most common co-morbid condition (102; 34.0%) followed by diabetes (80; 26.7%). The BSC provided to the patients included steroids (85.3%), haemostatics (89.7%), antivirals (91.3%), antibiotics (73.7%) and supplements (93.7%) ***(Suppl. Table 1)***.

The mean duration of COVID-19-related symptoms at enrolment was 8.41 days, and the most frequently reported COVID-19-related symptom was dyspnoea (92.7%), followed by cough (80.7%) and fever (71.3%).

### Primary outcome measures

#### i. Incidence of severe acute IRRs

Severe acute IRRs were reported in 1.3% of the patients; further details are presented under the safety section.

#### ii. a. One month mortality rate

A total of 20 deaths were reported in 300 patients (ITT population), resulting in a 1-month mortality rate of 6.7% [95% CI: 4%, 10%]. For the efficacy analysis population (mITT), a total of 18 deaths were reported, resulting in a mortality rate of 6.6% [18/287; 95% CI: 4%, 10%] at 1-month.

#### b. One month mortality: Sub-group analysis

Of the 222 patients at score-5 of the ordinal scale at baseline, there were 6 deaths (mortality rate 2.7%) and of the 78 patients at score-6 on the ordinal scale, there were 14 deaths (mortality rate 17.9%).

### Secondary outcome measures

#### i. Remission of respiratory symptoms

At the end of 30 days, 12 (4.0%) patients came off NIV after a mean duration of 5.6 days, and 275 (91.7%) patients were weaned-off from any supplemental oxygen therapy, with a mean duration of 8 days. (**Table 1**) There was no significant difference in time-to-independence from oxygen between NIV and any other oxygen therapy (p>0.05). Twelve (4.0%) patients moved to IMV, of which 2 (2.6%) survived by the end of extended follow-up of 90 days.

**Table 1:** Remission of respiratory symptoms.

| Statistics | Itolizumab; Full Analysis Set<br>(N=300) |  |
| --- | --- | --- |
|  | Time to independence from non-invasive mechanical ventilation (days) | Time to independence from oxygen therapy (days) |
| **n<sup>a</sup> or b (%)** | 12* (4.0) | 275** (91.7) |
| **Mean±SD** | 5.6±2.71 | 8.0±5.78 |
| **Median** | 6.0 | 6.0 |
| **Q1; Q3** | 3; 7.5 | 5; 9 |
| **Min; max** | 2; 10 | 1; 42 |

Note: N = Itolizumab number of patients in the relevant population, n<sup>a</sup> or n<sup>b</sup> = Number of patients on NIV or any supplemental oxygen, respectively, % = n<sup>a</sup> or b/N \*100. \*In addition, 18 (6%) patients either died while on NIV or discontinued due to an adverse event; so total of 12+18=30 patients. \*\*In addition, 25 (8.3%) patients either died while on O<sub>2</sub> therapy or discontinued due to an adverse event; so total of 275+25=300 patients. SD = standard deviation.

#### ii. Lung function determined by SpO_2_ and PFR

##### Stable/Improved SpO_2_

Stable SpO_2_ was defined as the absence of an increase in FiO_2_ to maintain SpO_2_ ≥92% and improvement of SpO_2_ was defined as a decrease in FiO_2_ to maintain SpO_2_ ≥92%. Most patients had stable or improved SpO_2_ without an increase in FiO_2_ by Day-7. Subsequently, for the rest of the visits, the same trend was followed **(Table 2)**.

**Table 2:** Stable/Improved SpO_2_ without increasing FiO_2_.

| Visit wise patients with stable/improved SpO<sub>2</sub> without increasing FiO<sub>2</sub><br>n (%) | Full Analysis Set: (N=300) |
| --- | --- |
| Day 7 | 259 (86.3) |
| Day 14 | 258 (86.0) * |
| Day 21 | 254 (84.7) * |
| End of study (Day 30) | 256 (85.3) * |
| *The status of patients who improved at Day 7 is carried forward to subsequent visits unless they subsequently worsened or died on study. |  |

##### PaO_2_/FiO_2_ ratio (PFR)

Mean PFR improved by about 50% by Day-7 (p<0.001). The increasing trend in PFR that was observed post Itolizumab dose was consistent till Day-30. Improvement from baseline was statistically significant at Days 3 and 7 (p<0.05) **(Figure 2)**.

**Figure 2:**
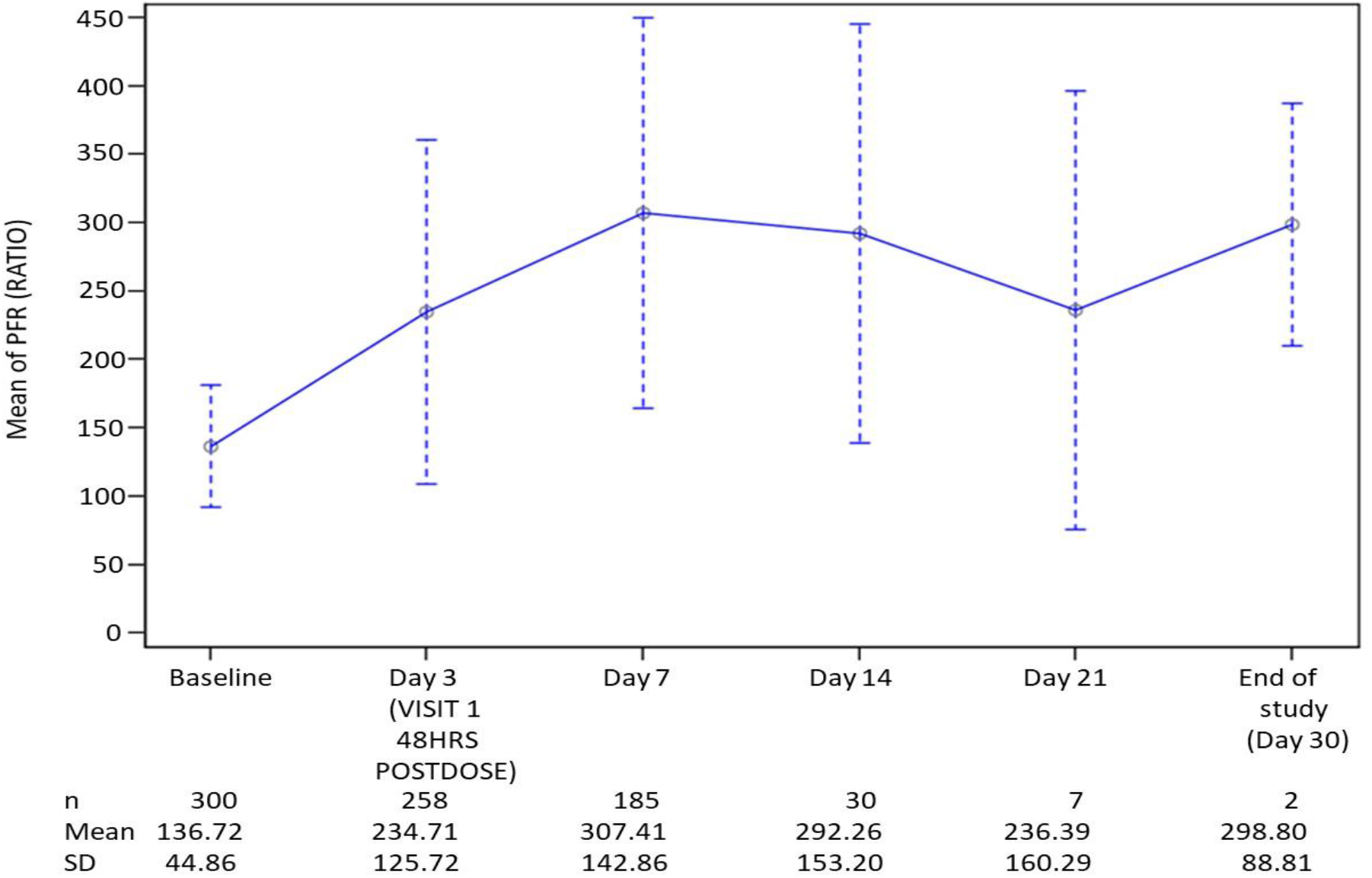
Summary of PaO_2_/FiO_2_ ratio (PFR) demonstrating improvement in the PFR that was consistent till Day 30.

#### iii. Duration of hospitalization

The summary of hospitalization duration, ICU stay and time to recovery by Day-30 are presented in **Table 3**. Overall, the patients took a median of 8 days for recovery and got discharged from the hospital by a median of 9.5 days. Patients who were severe required a median number of 11 days to get discharged from the ICU.

**Table 3:** Duration of hospitalization, ICU stay and time to recovery.

| Statistics | Full Analysis Set: (N=300) |  |  |
| --- | --- | --- | --- |
|  | Duration of hospitalization (days) during study* | Duration of ICU (days) during study* | Time to recovery (days) by Day 30 |
| **n (%)** | 284 (94.7) | 110 (36.7) | 255 (85) |
| **Mean±SD** | 10.9±5.38 | 13.0±7.22 | 9.80±6.49 |
| **Median** | 9.5 | 11 | 8 |
| **Min; max** | 3; 41 | 4; 41 | 7; 10 |
| Note: N = Itolizumab total number of patients, n = Number of patients in the relevant category, % = n/N *100<br>SD = standard deviation. Note 2: Time to recovery by Day 30 is defined as time to get to a score of 3 or below in the 8-point ordinal scale by Day 30.<br>*Duration of ICU stay has been presented and analysed only for 36.7% of patients requiring critical care (N=110). These patients further required additional stay in the ward before getting discharged. Duration of hospitalization is calculated for all irrespective of ICU admission needed or not (N = 284 [94.7%]). The median is expected to be different as patients in the ICU were more severe and required more stay in ICU as compared to other patients.<br>ICU = intensive care unit; SD = standard deviation. |  |  |  |

#### iv. Clinical Status as improvement of 1 or 2 points on the modified COVID-19 8-point ordinal scale

By Day-7, most patients started improving in the clinical status by at least 1 or more points on the ordinal scale. Two hundred and sixty (260; 86.7%) patients showed an improvement in the clinical status by at least 1-point on the 8-point ordinal scale and 26 (8.7%) showed worsening by at least 1-point on the scale. The rest either remained stable on their baseline ordinal score (9; 3.0%) or for few only the baseline data is available (5; 1.6%). Most of the patients were discharged by Day-30. Cumulatively, at Day-30, 260 (86.7%), 256 (85.3%), 132 (44.0%), 113 (37.6%) and 32 (10.7%) patients showed >1-point, >2-point, >3-point, >4-point and 5-point improvement on the 8-point ordinal scale, respectively. Patients who showed worsening of symptoms by >1 point, >2 points and 3-points were 26 (8.7%), 20 (6.7%) and 6 (2.0%), respectively ***(Suppl. Figure 2a&b)***.

#### v. Biomarkers (IL-6 and TNF-α)

A decreasing trend was observed from baseline to 48-hours post-infusion in mean IL-6 (227.8 ng/L to 161.9 ng/L; 28.9% decrease, p>0.42) and TNF-α levels (26.7 ng/L to 19.2 ng/L; 28.1%; p>0.16) **(Figure 3a & b)**. A similar decrease was observed from baseline to 48-hours post the second infusion.

**Figure 3a&b:**
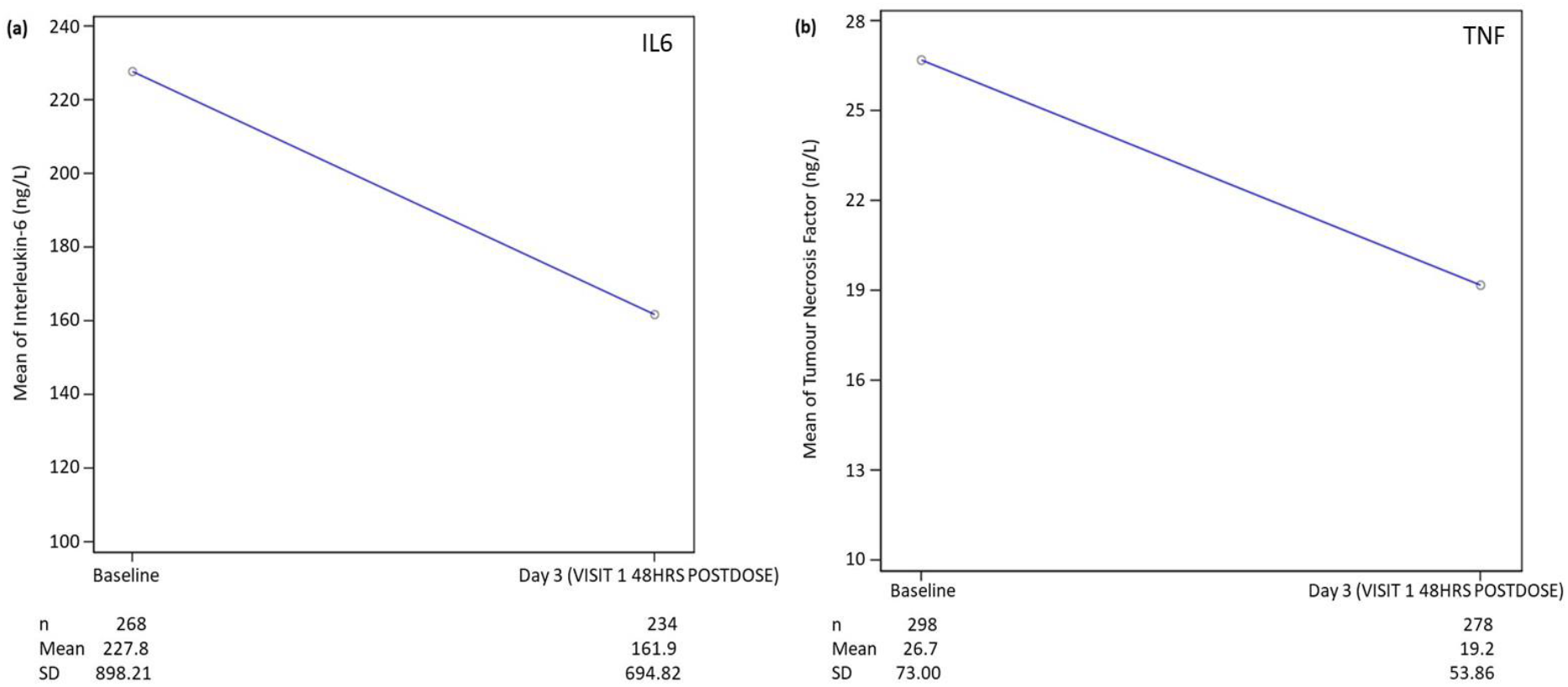
**a. Change in interleukin-6 and b: change in TNF-α levels post Itolizumab demonstrating a decreasing trend in levels from baseline to 48-hours post-dose. *Note: Normal values for IL-6 and TNF-α range from 0-7 ng/L and 4.6-12.4 ng/L, respectively*.**

#### vi. Change in inflammatory markers: CRP level, serum ferritin, D-dimer and LDH

A highly significant decrease from baseline was observed in the levels of CRP (p<0.0001; at Day-3 and Day-7), D-dimer (p<0.001; at Day-3 and Day-7), serum ferritin (p<0.01; at Day-7 and on the day of discharge) and LDH (p<0.001; at Day-7 and on the day of discharge) **(Figure 4 a,b,c,d)**.

**Figure 4a,b,c&d:**
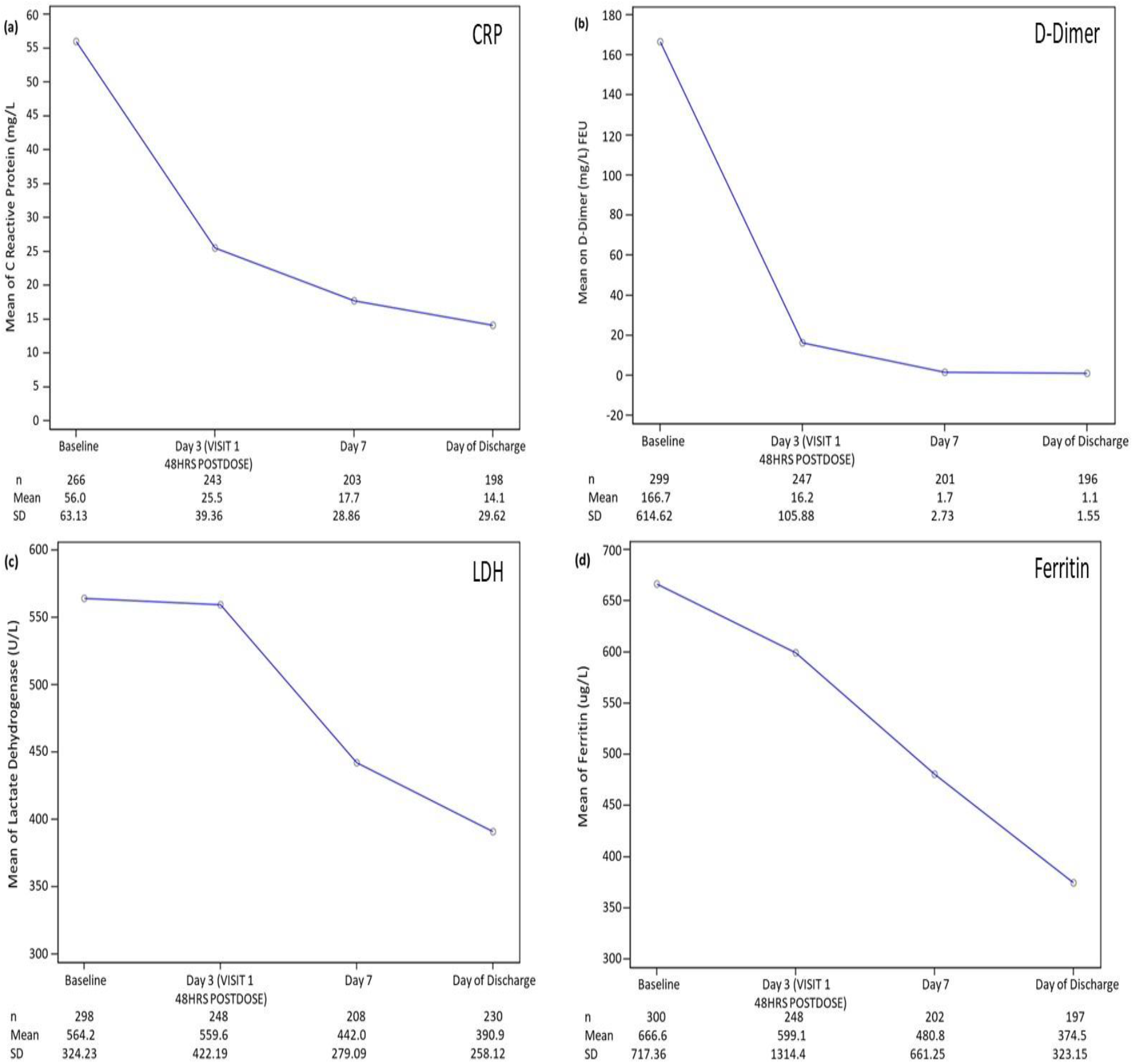
**a. Levels of C-reactive protein (CRP) post-Itolizumab first dose b. levels of D-Dimer post-Itolizumab first dose c. levels of lactate dehydrogenase (LDH) post-Itolizumab first dose and d. levels of ferritin post-Itolizumab first dose. All inflammatory markers decreased from baseline until day of discharge. *Note: Normal ranges of CRP, D-dimer, LDH and ferritin are 0-4.99 mg/L, 0-1 mg/L FEU, 134.97-214.16 U/L and 4.6-204 µg/L; respectively***.

#### vii. Safety

Itolizumab use was associated with an overall incidence of 21% TEAEs (123 TEAEs in 63 patients) till Day-30 of this study. The most commonly reported TEAEs were IRRs (7.3%) and lymphopenia (7%, including decreased lymphocyte count) **(Table 4)**. A total of 34 IRRs occurred in 22 patients of which 23 events were of CTCAE Grade-1, 5 events were of Grade-2, 4 events were of Grade-3 and 2 were of Grade-4. Twenty-one events of lymphopenia, including 2 events of decreased lymphocyte count, were reported till Day-30, and none of them were serious. Most of these events were of Grade-3 (19 events) and the majority (15 events) of these resolved before Day-30, with a median recovery time of 4 days. Thirty-eight TEAEs in 26 patients (8.7%) were drug related, of which 34 events were IRRs. Of the TEAEs, 30 were considered serious in 25 (8.3%) patients of which 5 events were of CTCAE Grade-4 and 20 events were of CTCAE Grade-5 (fatal) as reported by Day-30. The most frequently reported serious AEs (SAEs) were septic shock (6 events), worsening of ARDS (6 events) and respiratory failure (4 events). A total of 3 SAEs were considered to be related to Itolizumab, namely, bradycardia and decreased oxygen saturation, both reported by a single patient, and anaphylactic reaction reported in another patient, and all of these were considered as IRRs. The most common AEs by system organ class leading to death were respiratory disorders (N=8), cardiac disorders (N=6), infections (N=5) followed by renal disorders (N=1) and progression of the disease was considered as the underlying cause of these deaths. None of the TEAEs leading to death were related to Itolizumab. Overall, 12 TEAEs in 11 (3.7%) patients led to withdrawal from the study, and all TEAEs except one were IRRs. From Day-31 to Day-90, a total of 4 SAEs (bradycardia, dyspnoea, septic shock, all of them leading to death, and death by an unknown cause) were reported. None of them were related to Itolizumab.

**Table 4:**
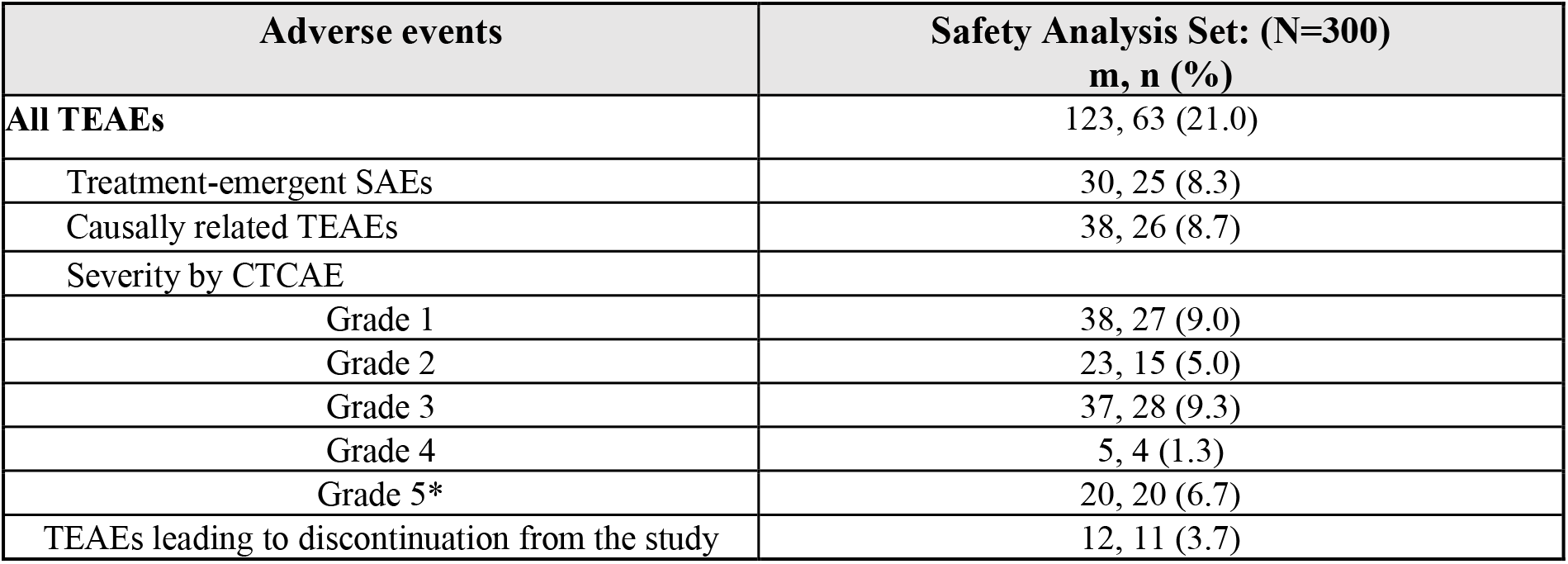
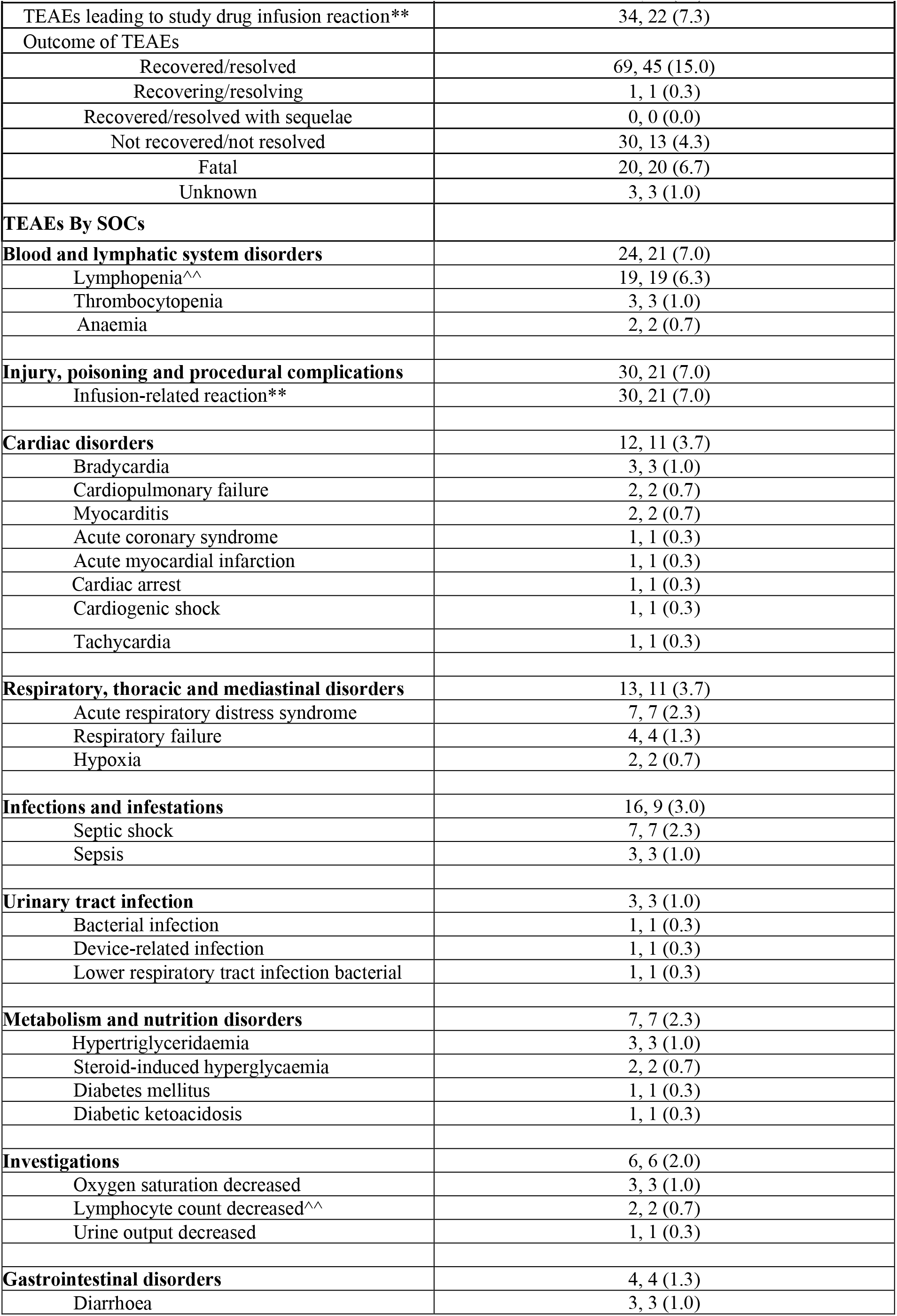
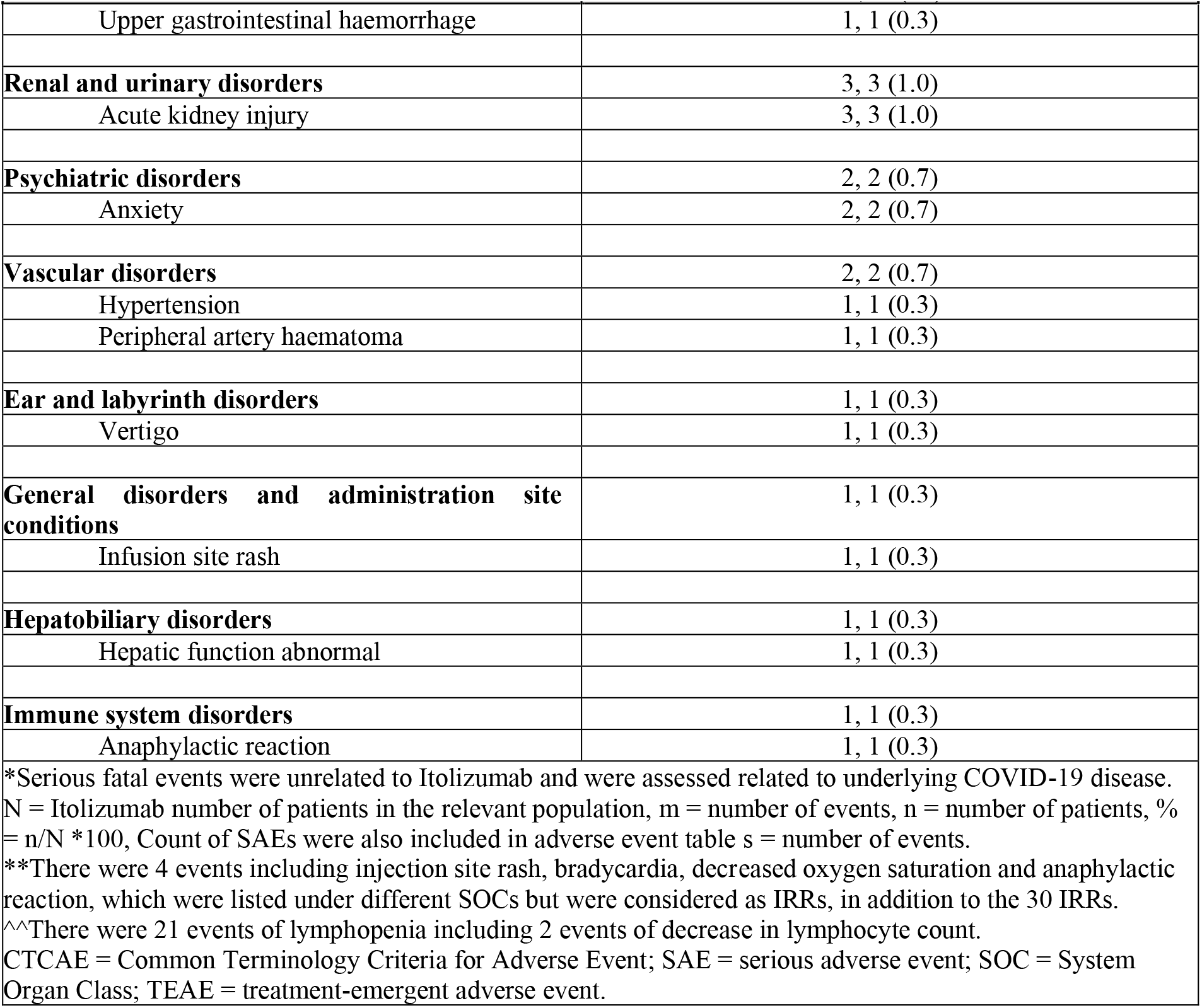
Summary of TEAEs.

| Adverse events | Safety Analysis Set: (N=300)<br>m, n (%) |
| --- | --- |
| All TEAEs | 123, 63 (21.0) |
| Treatment-emergent SAEs | 30, 25 (8.3) |
| Causally related TEAEs | 38, 26 (8.7) |
| Severity by CTCAE |  |
| Grade 1 | 38, 27 (9.0) |
| Grade 2 | 23, 15 (5.0) |
| Grade 3 | 37, 28 (9.3) |
| Grade 4 | 5, 4 (1.3) |
| Grade 5* | 20, 20 (6.7) |
| TEAEs leading to discontinuation from the study | 12, 11 (3.7) |

| **Adverse events** | **Safety Analysis Set: (N=300)**<br>**m, n (%)** |
| --- | --- |
| TEAEs leading to study drug infusion reaction** | 34, 22 (7.3) |
| Outcome of TEAEs |  |
| Recovered/resolved | 69, 45 (15.0) |
| Recovering/resolving | 1, 1 (0.3) |
| Recovered/resolved with sequelae | 0, 0 (0.0) |
| Not recovered/not resolved | 30, 13 (4.3) |
| Fatal | 20, 20 (6.7) |
| Unknown | 3, 3 (1.0) |
| **TEAEs By SOCs** |  |
| **Blood and lymphatic system disorders** | 24, 21 (7.0) |
| Lymphopenia^^ | 19, 19 (6.3) |
| Thrombocytopenia | 3, 3 (1.0) |
| Anaemia | 2, 2 (0.7) |
| **Injury, poisoning and procedural complications** | 30, 21 (7.0) |
| Infusion-related reaction** | 30, 21 (7.0) |
| **Cardiac disorders** | 12, 11 (3.7) |
| Bradycardia | 3, 3 (1.0) |
| Cardiopulmonary failure | 2, 2 (0.7) |
| Myocarditis | 2, 2 (0.7) |
| Acute coronary syndrome | 1, 1 (0.3) |
| Acute myocardial infarction | 1, 1 (0.3) |
| Cardiac arrest | 1, 1 (0.3) |
| Cardiogenic shock | 1, 1 (0.3) |
| Tachycardia | 1, 1 (0.3) |
| **Respiratory, thoracic and mediastinal disorders** | 13, 11 (3.7) |
| Acute respiratory distress syndrome | 7, 7 (2.3) |
| Respiratory failure | 4, 4 (1.3) |
| Hypoxia | 2, 2 (0.7) |
| **Infections and infestations** | 16, 9 (3.0) |
| Septic shock | 7, 7 (2.3) |
| Sepsis | 3, 3 (1.0) |
| **Urinary tract infection** | 3, 3 (1.0) |
| Bacterial infection | 1, 1 (0.3) |
| Device-related infection | 1, 1 (0.3) |
| Lower respiratory tract infection bacterial | 1, 1 (0.3) |
| **Metabolism and nutrition disorders** | 7, 7 (2.3) |
| Hypertriglyceridaemia | 3, 3 (1.0) |
| Steroid-induced hyperglycaemia | 2, 2 (0.7) |
| Diabetes mellitus | 1, 1 (0.3) |
| Diabetic ketoacidosis | 1, 1 (0.3) |
| **Investigations** | 6, 6 (2.0) |
| Oxygen saturation decreased | 3, 3 (1.0) |
| Lymphocyte count decreased^^ | 2, 2 (0.7) |
| Urine output decreased | 1, 1 (0.3) |
| **Gastrointestinal disorders** | 4, 4 (1.3) |
| Diarrhoea | 3, 3 (1.0) |
| Upper gastrointestinal haemorrhage | 1, 1 (0.3) |
| **Renal and urinary disorders** | 3, 3 (1.0) |
| Acute kidney injury | 3, 3 (1.0) |
| **Psychiatric disorders** | 2, 2 (0.7) |
| Anxiety | 2, 2 (0.7) |
| **Vascular disorders** | 2, 2 (0.7) |
| Hypertension | 1, 1 (0.3) |
| Peripheral artery haematoma | 1, 1 (0.3) |
| **Ear and labyrinth disorders** | 1, 1 (0.3) |
| Vertigo | 1, 1 (0.3) |
| **General disorders and administration site conditions** | 1, 1 (0.3) |
| Infusion site rash | 1, 1 (0.3) |
| **Hepatobiliary disorders** | 1, 1 (0.3) |
| Hepatic function abnormal | 1, 1 (0.3) |
| **Immune system disorders** | 1, 1 (0.3) |
| Anaphylactic reaction | 1, 1 (0.3) |
| *Serious fatal events were unrelated to Itolizumab and were assessed related to underlying COVID-19 disease. N = Itolizumab number of patients in the relevant population, m = number of events, n = number of patients, % = n/N *100, Count of SAEs were also included in adverse event table s = number of events.<br>**There were 4 events including injection site rash, bradycardia, decreased oxygen saturation and anaphylactic reaction, which were listed under different SOCs but were considered as IRRs, in addition to the 30 IRRs.<br>^^There were 21 events of lymphopenia including 2 events of decrease in lymphocyte count.<br>CTCAE = Common Terminology Criteria for Adverse Event; SAE = serious adverse event; SOC = System Organ Class; TEAE = treatment-emergent adverse event. |  |

### Infusion-Related Reactions

The incidence of severe acute IRRs was one of the primary outcome measures. Of the 34 drug-related IRRs, 6 events in 4 patients (1.3%) were of Severity Grade-3 and −4 and 3 IRRs were reported as SAEs. All IRRs were treated as per the institutional IRR treatment protocol and were considered resolved/recovered on the same day. IRRs led to the early discontinuation of 10 patients from the study. Incidence of severe, acute IRRs and higher, causally related to Itolizumab, is presented in **Table 5**.

**Table 5:** Treatment-emergent severe acute infusion-related reactions causally related to Itolizumab.

| Description | Itolizumab<br>(N=300)<br>m, n (%) |
| --- | --- |
| Number of patients with at least one related infusion-related reactions any grade causally related to Itolizumab | 34, 22 (7.3) |
| Number of patients with at least one severe acute infusion-related reactions and higher causally related to Itolizumab | 6, 4 (1.3) |
| N = Number of patients in the relevant population, m = number of events, n = number of patients with IRRs, % = n/N *100.<br>IRR = infusion-related reaction. |  |

## Discussion

The inflammatory pathophysiology of COVID-19 has encouraged research on several immunomodulatory treatments in moderate-to-severe cases of COVID-19, but the results have been inconclusive. A meta-analysis of eight trials on the use of Tocilizumab has shown no significant difference in 28-day mortality barring one trial (REMAP-CAP) in critically ill patients.^25^ In the current study, the 1-month mortality rate for patients, who received one full infusion of Itolizumab, was observed to be 6.7%. A meta-analysis done on six concurrently running COVID-19 trials in India with 11,852 hospitalized patients, representing a similar patient population receiving standard of care, showed an average mortality rate of 27% with 95% CI [16% to 42%] in the control-arm ***(Suppl. Figure 3, Suppl. Table 5)***.^26-31^ Mortality at 1-month has been observed to be 19.4%, 35% and 15.2% in the control-arms and 19.7%, 31% and 11.4% in the treatment-arms of the COVACTA, RECOVERY (Tocilizumab) and ACTT-1 trials, respectively.^24,25,32^ A lower percentage of patients received systemic corticosteroids in the COVACTA (tocilizumab, 33.7%; control, 52.1%) and ACTT-1 (Remdesivir, 21.6%; control, 24.4%) trials though a higher proportion of patients received steroids in the RECOVERY trial (tocilizumab, 82%; control, 82%) and in the current study (Itolizumab, 85.3%). In the RECOVERY and COVACTA tocilizumab treated arms, patients who required any supplemental oxygen (equivalent to score-5 on the COVID-19 8-point ordinal scale) had mortality rates of 19% and 11.5% and those requiring NIV/HFNC (equivalent to score-6 on the ordinal scale) had mortality rates of 38% and 19.1%, respectively.^24,31^ In the current study, mortality was 2.7% for ordinal score-5 and 17.9% for ordinal score-6 patients treated with Itolizumab.

In our study, 91.7% of patients got weaned off from any kind of supplemental oxygen within 6 (3-7.5) days. Only 4% of patients progressed to IMV of which 2.6% did not survive till Day-30. At the end of Day-30, majority of the patients (86.7%) showed an improvement on the ordinal scale by 1 or more points from the baseline score of 5/6 and 8.7% patients showed worsening of symptoms by 1 or more points on the ordinal scale. Itolizumab’s intervention, thus, can be helpful before the patient progresses to severe ARDS requiring IMV. Median time to recovery with Itolizumab was 8 (7-10) days and it was 10 (9-11) days with Remdesivir in the ACTT-1 trial, 20 (17-27) days with Tocilizumab in the COVACTA trial and 19 days with Tocilizumab in the RECOVERY trial.^24,25,32^ Taken together, the mortality, recovery and remission observations with Itolizumab suggest a survival as well as an early recovery benefit.

Progression of COVID-19 complications is phasic and there is an unmet need to provide the right therapy, beyond steroids, to the right patients just as the systemic hyperinflammation sets in and quickly progresses towards a massive release of cytokines. Hypoxic respiratory failure in patients with COVID-19 is associated with high levels of pro-inflammatory cytokines, such as IL-6, and TNF-α, and elevated concentrations of D-dimer, ferritin and CRP,^25^ and these need to be closely monitored as prognostic markers for potential progression to critical illness.^33,34^ In this study, the biomarkers, IL-6 and TNF-α, which got elevated with the progression of the disease, showed a decreasing trend and the increased inflammatory markers (LDH, Ferritin, D-Dimer and CRP) decreased significantly by the time of discharge. The resolution of high levels of inflammatory markers also correlated with an improvement of lung function. Patients treated with Itolizumab continued to maintain stable/improved SpO_2_ without increasing FiO_2_ till Day-30 and showed an increasing trend in PFR, which was consistent till the day of discharge. Since Itolizumab can effectively change the hyperinflammation milieu by arresting the inflammatory cytokine release from T-cells, its use holds promise in other conditions such as viral haemorrhagic fevers, SARS, MERS and post-CART-mediated cytokine release syndrome.

No new safety concerns were observed in this study. The most frequently reported TEAEs, namely, IRRs (7.3%) and lymphopenia (7%; including decreased lymphocyte count), are typically characteristic of treatment with monoclonal antibodies.^9,15,35^ All IRRs resolved on the same day and most of the lymphopenia events were transient and resolved before Day-30, with a median recovery time of 4 days. It has been observed that on increasing the infusion time from 2 hours to 5-6 hours, the incidence of IRRs can be reduced.^9^ In the current study, there were 1.3% IRRs reported as Grade 3/4 and 3.3% of patients withdrew from the study due to an IRR. Of the 30 SAEs observed, none of the 20 fatal SAEs were related to the drug. The most common SAEs were septic shock, worsening of ARDS and respiratory failure, which are generally observed in patient populations with co-morbid conditions.^36,37^ The three SAEs related to Itolizumab, namely, bradycardia, anaphylactic reaction and decreased oxygen saturation, were considered as IRRs. The incidence of septic shock in this study (2%) is similar to other reports in COVID-19 patients, where it ranges from 2-4%.^36-39^ In this study, there was no increase in incidences of serious infections or septicaemia and no new infections, such as mucormycosis, were observed when compared to previous clinical experiences or concurrent data.

Overall, the clinical trial data suggest that Itolizumab is generally safe and well-tolerated. A subcutaneous version of Itolizumab under development has an added advantage to be used as an outpatient intervention and holds a promise to offset the IRRs. This trial has the limitation of being a single-arm, open-label study. Typically, collection of concurrent data from the same hospitals/centres can be useful for comparison, however, amidst a pandemic setting this was not feasible. The meta-analysis on concurrent control data collected from nearly 12,000 patients during the same period has the limitation of having high heterogenicity, though, enough care was taken to match the populations, disease severity, BSC and treatment with other immunomodulators.

## Conclusions

Itolizumab is well tolerated and shows no new safety concerns. Reduction in the inflammatory markers, consistent with improvement in lung function and the COVID-19 ordinal scores, underscores the role of early and timely intervention with Itolizumab in controlling the COVID-19 inflammatory phenotype. Itolizumab treatment indicates a survival and recovery benefit at 1-month, acting through its unique mechanism of action of inhibiting the T-cell mediated inflammatory milieu, in COVID-19 patients hospitalized with ARDS.

## Sponsor

The study was funded by Biocon Biologics Limited, and the sponsors did not have any role in patient recruitment and management.

Sandeep N. Athalye, as the guarantor of this work, takes full responsibility for the work, including the study design, access to data and the decision to submit and publish the manuscript.

## Data Availability

All data produced in the present study are available upon reasonable request to the authors

## Author contributions

RD, SL, SD, RA, AM, SV and SNA contributed to the conception and design of the work. RKR, CR, RD, BKT, SD and RA contributed substantially to the conduct of this study, acquisition of the data and interpretation of the results. SNA is the guarantor of the study. AM contributed to the statistical analyses, meta-analysis and interpretation of data. RA was responsible for medical monitoring and NMC was responsible for supervision and acquisition of safety data and review and analysis of the SAE/AEs. All authors have critically reviewed the manuscript for important intellectual content and approved the same for publication.

## Acknowledgement

Medical writing support was provided by Shivani Mittra, PhD (Biocon Biologics Limited). Dev B Baruah, M. Pharm and Vathsala Jayanth, M.D. provided editorial support (both from Biocon Biologics Limited). Statistical Programming support was provided by Bhatu K Patil and Avinash Sevankar, M.Sc. (both from Biocon Biologics Limited).

