## Supplementary figures and images for "REcovery and SURvival of patients with moderate to severe acute REspiratory distress syndrome (ARDS) due to COVID-19: a multicentre, single-arm, Phase IV Itolizumab Trial: RESURRECT"

### Suppl Figure 1

#### Slide 1
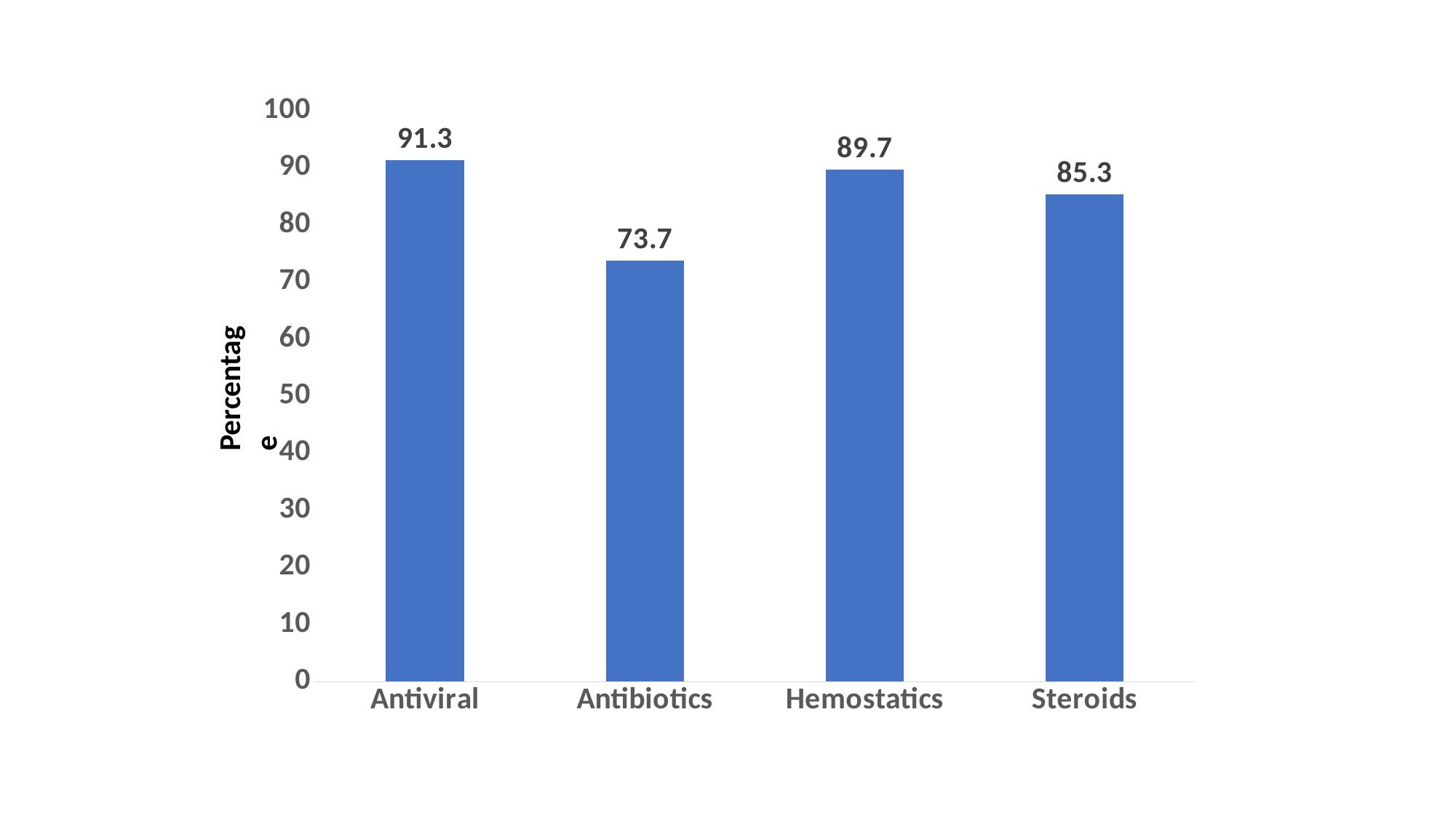

##### Chart
| Category | Series 1 |
|---|---|
| Antiviral | 91.3 |
| Antibiotics | 73.7 |
| Hemostatics | 89.7 |
| Steroids | 85.3 |Percentage

### Suppl Figure 3

#### Slide 1
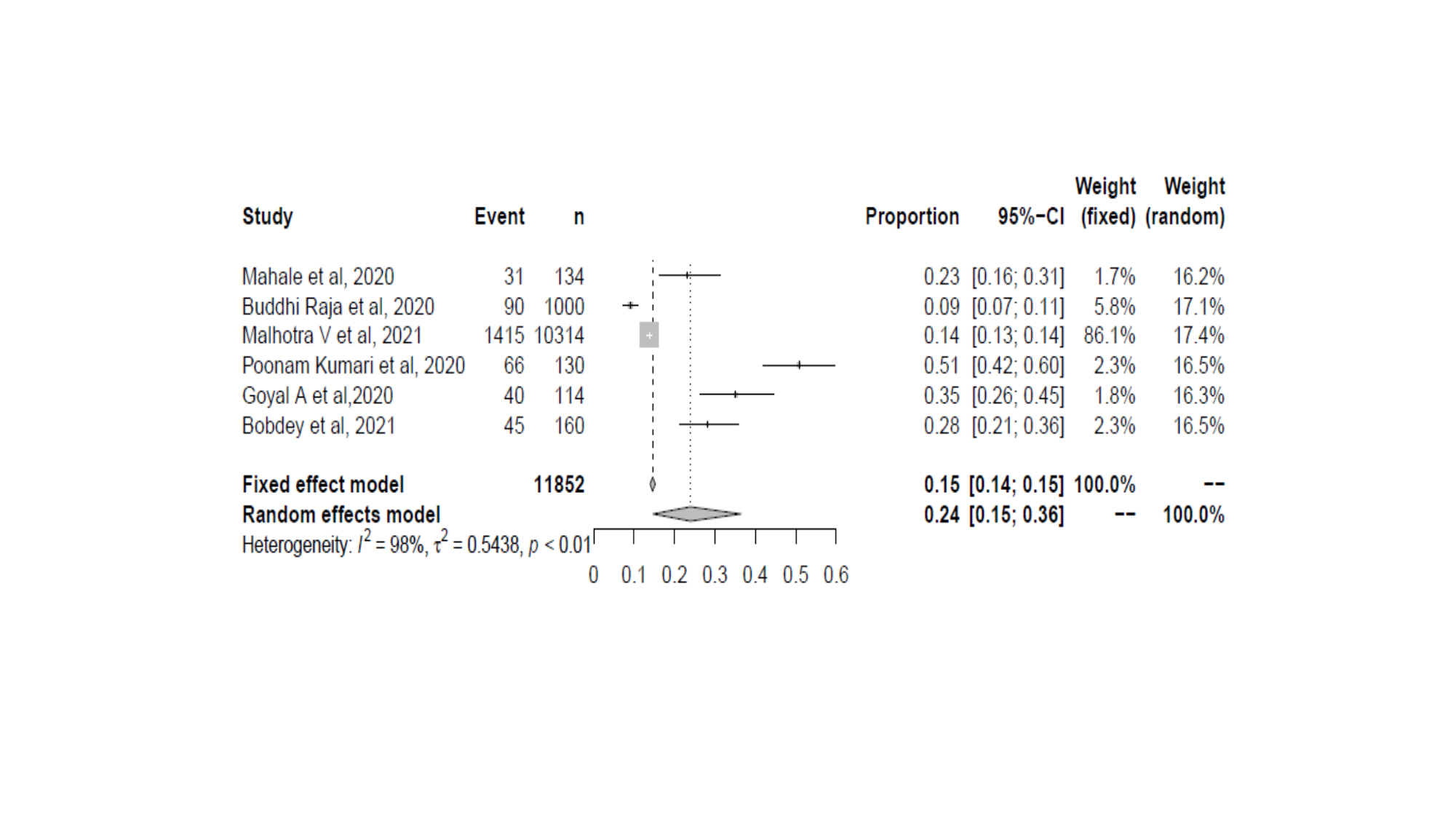
