## Supplementary material for "REcovery and SURvival of patients with moderate to severe acute REspiratory distress syndrome (ARDS) due to COVID-19: a multicentre, single-arm, Phase IV Itolizumab Trial: RESURRECT": Suppl Figure 2

### Slide 1
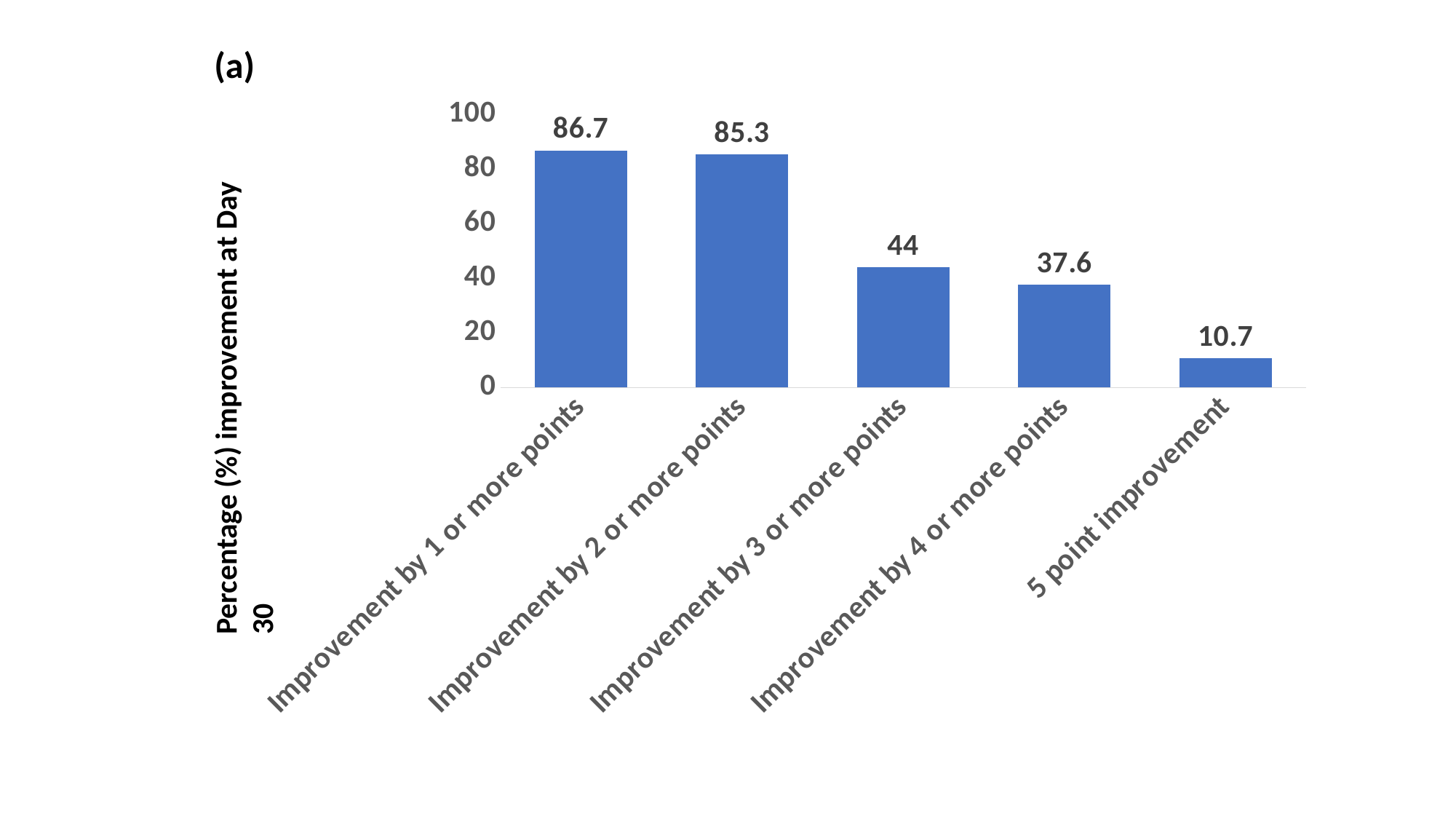

(a)
#### Chart
| Category | Series 1 |
|---|---|
| Improvement by 1 or more points | 86.7 |
| Improvement by 2 or more points | 85.3 |
| Improvement by 3 or more points | 44.0 |
| Improvement by 4 or more points | 37.6 |
| 5 point improvement | 10.7 |Percentage (%) improvement at Day 30

### Slide 2
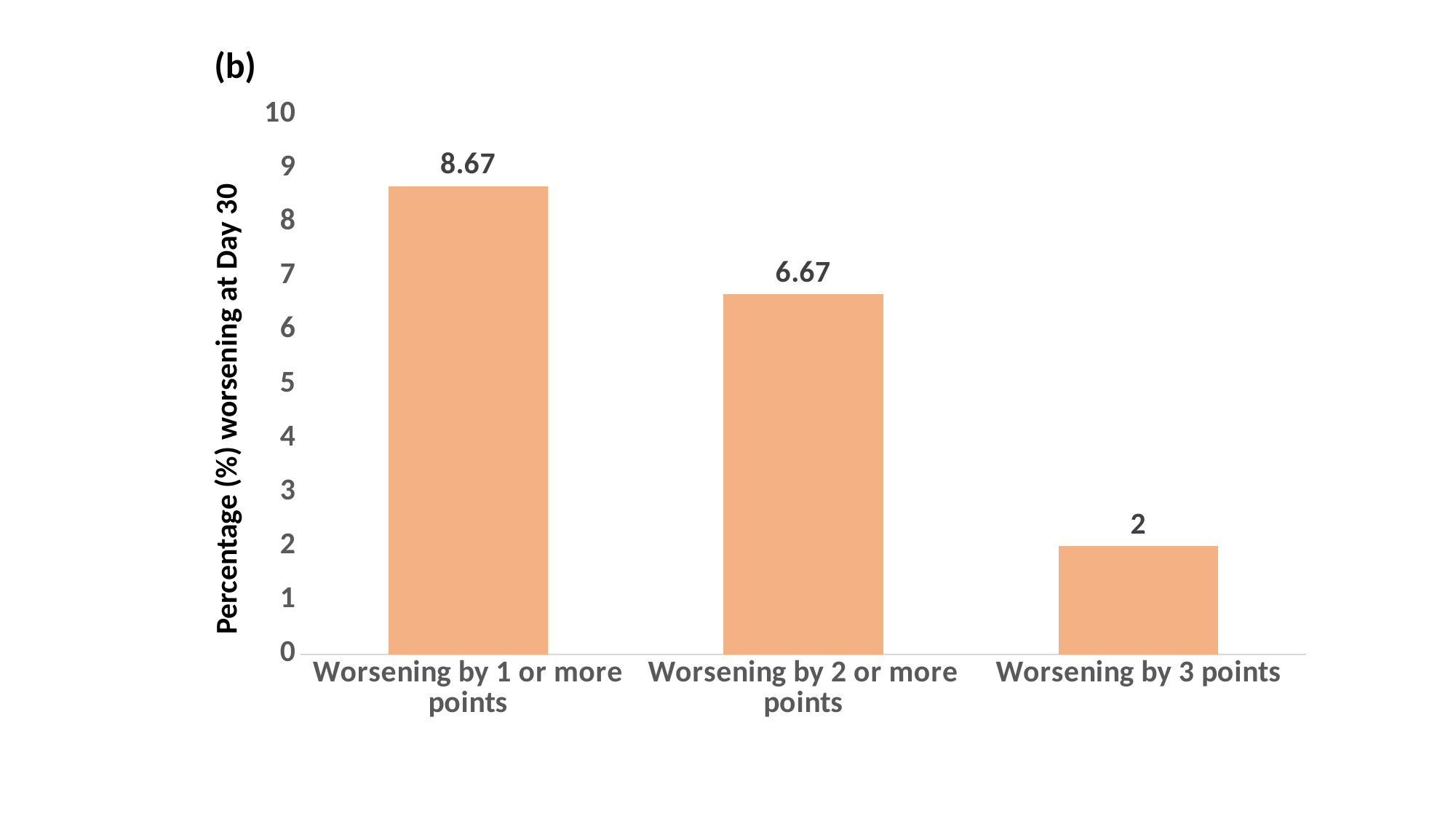

(b)
#### Chart
| Category | Series 1 |
|---|---|
| Worsening by 1 or more points | 8.67 |
| Worsening by 2 or more points | 6.67 |
| Worsening by 3 points | 2.0 |Percentage (%) worsening at Day 30
