## Supplementary appendix for "REcovery and SURvival of patients with moderate to severe acute REspiratory distress syndrome (ARDS) due to COVID-19: a multicentre, single-arm, Phase IV Itolizumab Trial: RESURRECT"

**Assessments**

Vitals, including pulse, blood pressure, respiratory rate and temperature (highest temperature of the day), were recorded daily. Hematology assessments including hemoglobin, total leukocyte count, differential leukocyte count, absolute neutrophil count, absolute lymphocyte count and platelet count were done at screening, baseline, 48 hours post first dose infusion, at Day-7, weekly (± 1 day) until discharge and at the day of discharge. Hematology assessments were performed additionally at 48 hours post second dose in patients receiving the second dose of Itolizumab. Blood samples for biomarker (IL-6 and TNF-α) assessments were collected at pre-dose (screening/baseline) and after 24-48 hours post each infusion. Blood samples for assessments of inflammatory markers (CRP, serum ferritin, D-dimer and LDH) were collected at screening/baseline, 48 hours post first dose infusion, at Day-7 and then weekly until discharge. SPO_2_ (highest stable value for approximately 1 hour of the day or as per the Principal Investigator’s discretion) and corresponding FiO_2_ were recorded daily until discharge. PFR and sequential organ failure assessment (SOFA) score were recorded at screening, baseline, approximately at 48 hours post-dose, at Day-7 (± 1 day) and on the day of discharge.
