## Supplementary Tables for "REcovery and SURvival of patients with moderate to severe acute REspiratory distress syndrome (ARDS) due to COVID-19: a multicentre, single-arm, Phase IV Itolizumab Trial: RESURRECT"

**Suppl. Table 1: Oxygen delivery mode at baseline**

| **Oxygen delivery mode at baseline** |  |
| --- | --- |
| **Baseline demographic variables** | **Itolizumab**  **(N=300)** |
| Invasive mechanical ventilation | 0 (0.0) |
| CPAP (Non-invasive ventilation mask) | 0 (0.0) |
| BiPAP (Non-invasive ventilation mask) | 30 (10.0) |
| Venturi mask | 0 (0.0) |
| High flow nasal cannula | 48 (16.0) |
| Non-rebreather mask | 74 (24.7) |
| Simple face mask | 105 (35.0) |
| Nasal prongs | 43 (14.3) |
| BiPAP = bi-level positive airway pressure; CPAP = continuous positive airway pressure. | |

**Suppl. Table 2: The modified COVID-19 8-point ordinal scale at baseline**

| **The modified COVID-19 8-point ordinal scale at baseline** |  | |
| --- | --- | --- |
| **Baseline oxygen requirement** | **Itolizumab**  **(N=300)**  n (%) | |
| 1. Not hospitalized; no limitations of activities | 0 (0.0) | |
| 2. Not hospitalized; limitation of activities; home oxygen requirement; or both | 0 (0.0) | |
| 3. Hospitalized; not requiring supplemental oxygen and no longer requiring ongoing medical care | 0 (0.0) | |
| 4. Hospitalized; not requiring supplemental oxygen but requiring ongoing medical care | 0 (0.0) | |
| 5. Hospitalized, requiring any supplemental oxygen | 222 (74.0) | |
| 6. Hospitalized; requiring non-invasive ventilation or use of high-flow oxygen devices | 78 (26.0) | |
| 7. Hospitalized, receiving invasive mechanical ventilation or extracorporeal membrane oxygenation (ECMO) | 0 (0.0) | |
| 8. Death | | 0 (0.0) |
| n = number of patients. | | |

**Suppl. Table 3: Itolizumab dosing details**

| **Number of Itolizumab doses provided to patients** | |
| --- | --- |
| **Description** | **Itolizumab**  **(N=300)**  **n (%)** |
| Number of patients received Itolizumab | 111 (37.0) |
| Number of patients received lyophilized Itolizumab | 189 (63.0) |
| Number of patients with infusion interruption | 13 (4.3) |
| **Reason for interruption** |  |
| Adverse event | 13 (4.3) |
| Number of patients with two doses of Itolizumab |  |
| Yes | 9 (3.0) |
| No | 287 (95.7) |
| n = number of patients. | |

**Suppl. Table 4.** **Participant disposition, demographics, and baseline characteristics**

|  | **Variable** | **Itolizumab**  **(N=300)**  **n (%)** | |
| --- | --- | --- | --- |
|  | FAS population* | 300 (100) | |
|  | Efficacy population** | 287 (95.7) | |
|  | Safety population*** | 300 (100) | |
|  | Number of completed patients at Day 30 | 256 (85.3) | |
|  | Discontinued | 44 (14.7) | |
|  | Reasons for discontinuation  Death^#^  Adverse event  Patient/guardian decision | 18 (6.0)  11 (3.7)  15 (5.0) | |
|  | **Demographic and baseline characteristics** | | |
|  | Age (years) | |  |
| N | | | 300 (100) |
| Mean (SD) | | | 53.3 (14.9) |
| Sex | | |  |
| Female | | | 76 (25.3) |
| Male | | | 224 (74.7) |
|  | Race | |  |
|  | Asian | | 300 (100) |
|  | D-Dimer (mg/L FEU)  Mean (SD) | | 166.7 (± 614.62) |
|  | D-Dimer (mg/L FEU)  Median (range) | | 1.0 (0.005; 6425.6) |
|  | Ferritin (µg/L)  Mean (SD) | | 666.6 (± 717.36) |
|  | Ferritin (µg/L)  Median (range) | | 498.6 (21.6; 6800) |
|  | LDH (U/L)  Mean (SD) | | 564.21 (± 324.23) |
|  | LDH (U/L)  Median (range) | | 8.6 (1.7535; 45.3405) |
|  | C-Reactive protein (mg/L)  Mean (SD) | | 56.0 (± 63.13) |
|  | C-Reactive protein (mg/L)  Median (range) | | 35.0 (0.6; 315.98) |
|  | Lymphocyte (cells/mm^3^)  Mean (SD) | | 1482.7 (± 1129.5) |
|  | ^*^Full Analysis Set Included all patients who are enrolled in the study; **Efficacy Analysis Set: Included all patients of the FAS population who received at least one full infusion of Itolizumab (without any major protocol deviation; decided before the database lock); ^***^Safety Analysis Set: Included all patients who received a partial or complete infusion of Itolizumab and were used for safety endpoint analysis; ^#^Two patients who were discontinued from the treatment due to AEs died later; n=number of patients. AE = adverse event; FAS = Full Analysis Set; LDH = lactate dehydrogenase; SD = standard deviation. | | |

**Suppl. Table 5: Historical mortality, recovery rates in control groups along with key demographics and baseline characteristics in Indian hospitals**

| **Parameters** | **Buddhiraja et al 2020** | **Malhotra et al 2021** | **Mahale et al 2020** | **Kumari et al 2021** | **Goyal at al 2020** | **Bobdey et al 2021** |
| --- | --- | --- | --- | --- | --- | --- |
| **Study type** | Record based, observational study | Retrospective cohort  study | Retrospective cohort study | Off-label /Compassionate use study | Chart review | Retrospective study |
| **Sample size** | 976 | 10,314 | 134 | 150 | 114 | 1233 |
| **Treatment arms** | NA | NA | NA | Itolizumab (n=20) vs Control (n=130) | NA | NA |
| **Sex (% of M/F patients)** | 67.1/32.9 | 58.50/ 41.50 | 68.0/32.0 | NAV | 78.1/21.9 | 79.8/20.2 |
| **Age (Mean/ Median in Years)** | 47.5 | 46.43 | 55.6 | NAV | 58.2 | 41.63 |
| **PFR at entry** | NAV | NAV | <300 for 60% patients | <200 | <300 | NAV |
| **Disease severity** | Mild to Severe | Moderate to Severe | Mild to Severe | Moderate to Severe | Moderate to Severe | Mild to Severe |
| **Presence of at least one Comorbidity (%)** | 18.24 | NAV | 72.0 | NAV | 51.8^α^ | 5.7 |
| **SoC and concomitant medications** | HCQ+ azithromycin, Ivermectin, Steroids, Tocilizumab, Convalescent Plasma Therapy | Remdesivir, Steroids, Ivermectin, Tocilizumab, HCQ and anticoagulants | Tocilizumab, colchicine, HCQ, methylprednisolone, etoricoxib, antibiotics | NAV | Steroid therapy and antibiotics few received antivirals/tocilizumab | NAV |
| **Mortality Rate (%)** | 10.5^β^ | 13.72 | 26.9 | 35 vs 60^β^ | 43.85 | 5.8 |
| **Recovery (%)** | 89.45 | 86.28 | 68.7 | 65 vs 40 | 56.14 | 94.16 |

**^α^Derived from diabetes medical history**

**^β^1-Month mortality rate**

***Note: To account for the possibility of different effect sizes across studies, a random effects meta-analysis model was used to assess heterogeneity of effects across studies using the I^2^ statistic (I^2^ =98%, p<0.01) in the R Studio software.***
